# Tracking seizure cycles beats a prospective moving average

**DOI:** 10.1101/2025.11.03.25338700

**Authors:** Rachel E. Stirling, Benjamin H. Brinkmann, Dean R. Freestone, Philippa J. Karoly

## Abstract

This commentary addresses the debate regarding the predictive value of multiday seizure cycles versus simple statistical baselines. Multidien seizure cyclicity is a prevalent, patient-specific phenomenon with promise for epilepsy management. We challenge the assertion that cycle tracking is no better than a 90-day moving average, which is an inherently retrospective model that lags changes in seizure likelihood. We compared a causal cyclic forecast to a prospectively applied moving average across a large seizure diary cohort (n=768) and two gold-standard chronic EEG cohorts (n=24). At the group level for the EEG and diary cohorts, cycle tracking demonstrated significantly superior accuracy to the moving average for both hourly and daily forecasts (p < 0.0001). These results confirm that event-based cyclical models offer more accurate, simulated real-world forecasts. We conclude that robust forecasting tools must prioritize the detection and modeling of seizure cycles to move beyond simple baseline performance and provide actionable clinical utility.

---

Multidien cycles of seizures have emerged as a prevalent phenomenon in epilepsy, alongside well documented circadian patterns. These longer rhythms have been observed at individual-specific weekly, monthly, seasonal, or other multiday periods and occur across sex, medications, age, and epilepsy type (Karoly et al., 2021). Detecting and tracking individuals’ seizure cycles holds promise for epilepsy management, and significant advances have been made in the reliable extraction of seizure cycles from different data sources. Projecting cycles forward in time to forecast future periods of seizure risk has been demonstrated successfully, although several challenges remain. Chang and colleagues (Chang et al., 2025) contend that the forward projection of multiday seizure cycles is no better than using the historic (90-day) long-term average of seizure rates to estimate the chance of a seizure the next day. On an individual level, this will be true for some people and not others. It is likely that the 90-day historic average provides an approximation of longer (i.e. monthly) rhythms and clustered seizures, although it is likely less precise for people with shorter (i.e. weekly) rhythms and certainly contains no information about circadian trends. While benchmarking seizure forecasts against a historic moving average is a reasonable baseline, suggesting that tracking seizure cycles performs no better than tracking a moving average rate in the general population is not currently supported by data. We were unable to replicate the results reported for the 90-day moving average method.

We compared a prospectively applied 90-day moving average forecast to a causal cyclic forecast (Nasseri et al., 2025) in a large seizure diary cohort (n=768) (Stirling et al., 2024) and two small, gold-standard benchmarked chronic EEG cohorts: the My Seizure Gauge (MSG) cohort, consisting of 11 subjects who had a chronic EEG monitoring device (UNEEG SubQ, NeuroPace RNS, or Medtronic Summit RC□+□S) for 6-24 months (Nasseri et al., 2025); and the NeuroVista (NV) cohort, consisting of 13 subjects who were implanted with the NeuroVista device for 6-24 months (Cook et al., 2013). In the chronic EEG cohort, cycle tracking was more accurate than a moving average in all but one of the MSG and all but one of the NV cohorts, for both hourly and daily pseudo-prospective forecasts (with forecast horizons extending to the next seizure event). Cycles tracking performed significantly better than a moving average at the group level comparing both hourly and daily AUC (p < 0.0001, paired t-test, combined MSG and NV cohorts). In the seizure diary cohort, cycle tracking was more accurate than 86% and 63% of moving average forecasts at the hourly and daily timescales, respectively. These proportions are similar to the number of people with significant circadian and multiday cycles (∼90% and ∼60%) observed in longitudinal seizure diary cohorts (Karoly et al., 2018; Leguia et al., 2021). Hourly and daily group level performance confirmed cycles tracking was significantly better than moving average (p < 0.0001, paired t-test). Results are shown in Table 1 and both chronic EEG datasets are publicly available (MSG Cohort: https://www.epilepsyecosystem.org/my-seizure-gauge-1 and NV Cohort: https://doi.org/10.26188/5b6a999fa2316). These comparisons suggest that event-based seizure cycles provide significantly more accurate forecasts than a moving average in a simulated real-world setting, highlighting the importance of prospective implementation during testing. Moving averages, though simple, are inherently retrospective, as they estimate risk from recent seizure counts within a fixed window. This approach, which may effectively track long cycles retrospectively, lags impending changes in seizure probability, depends heavily on window length, and is unable to project an underlying cycle forward in time. As a result, it performs poorly at forecasting lead seizures following quiescent periods and is less reliable than causal event-based cyclical models.

**Table 1.**
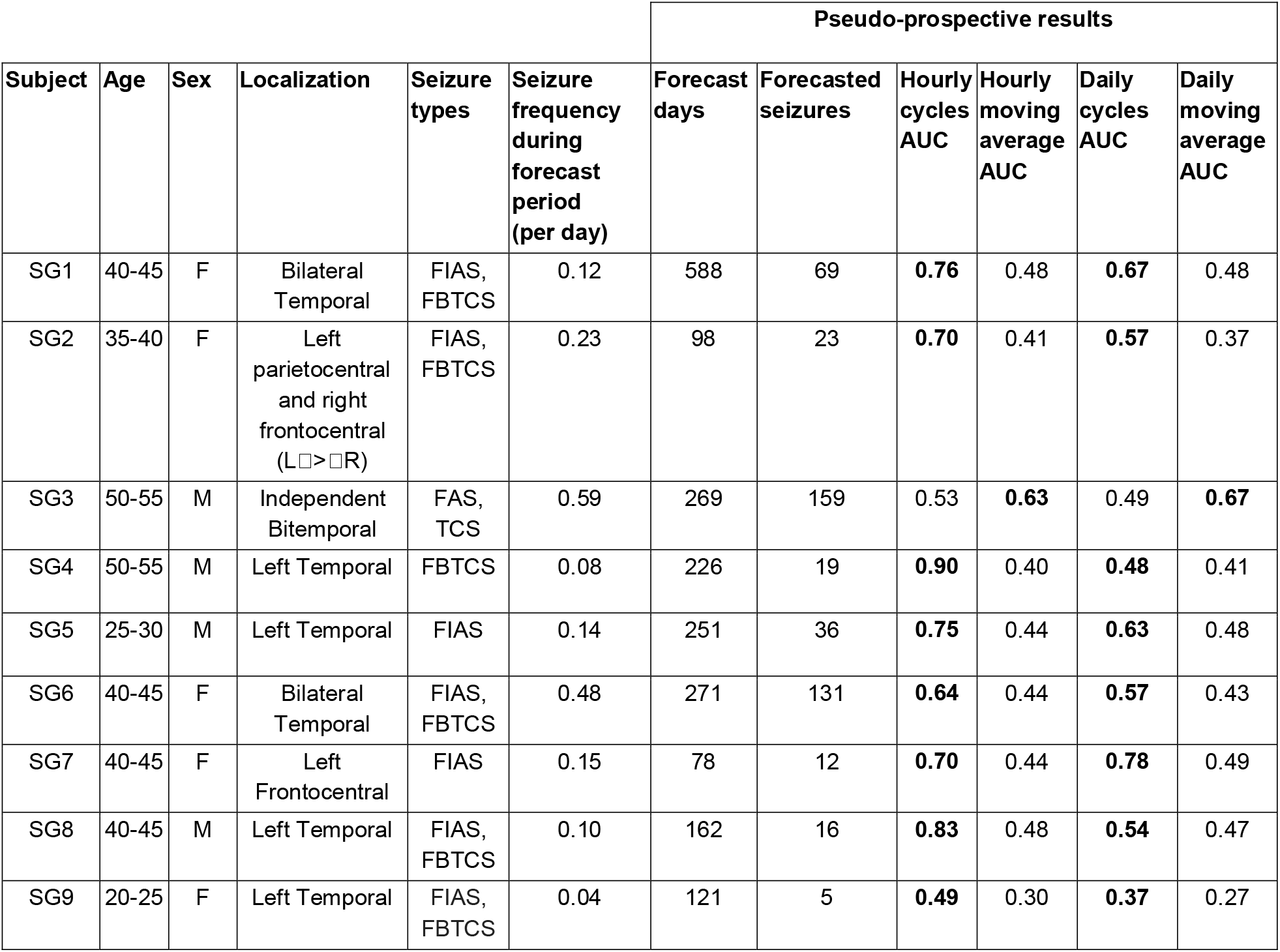
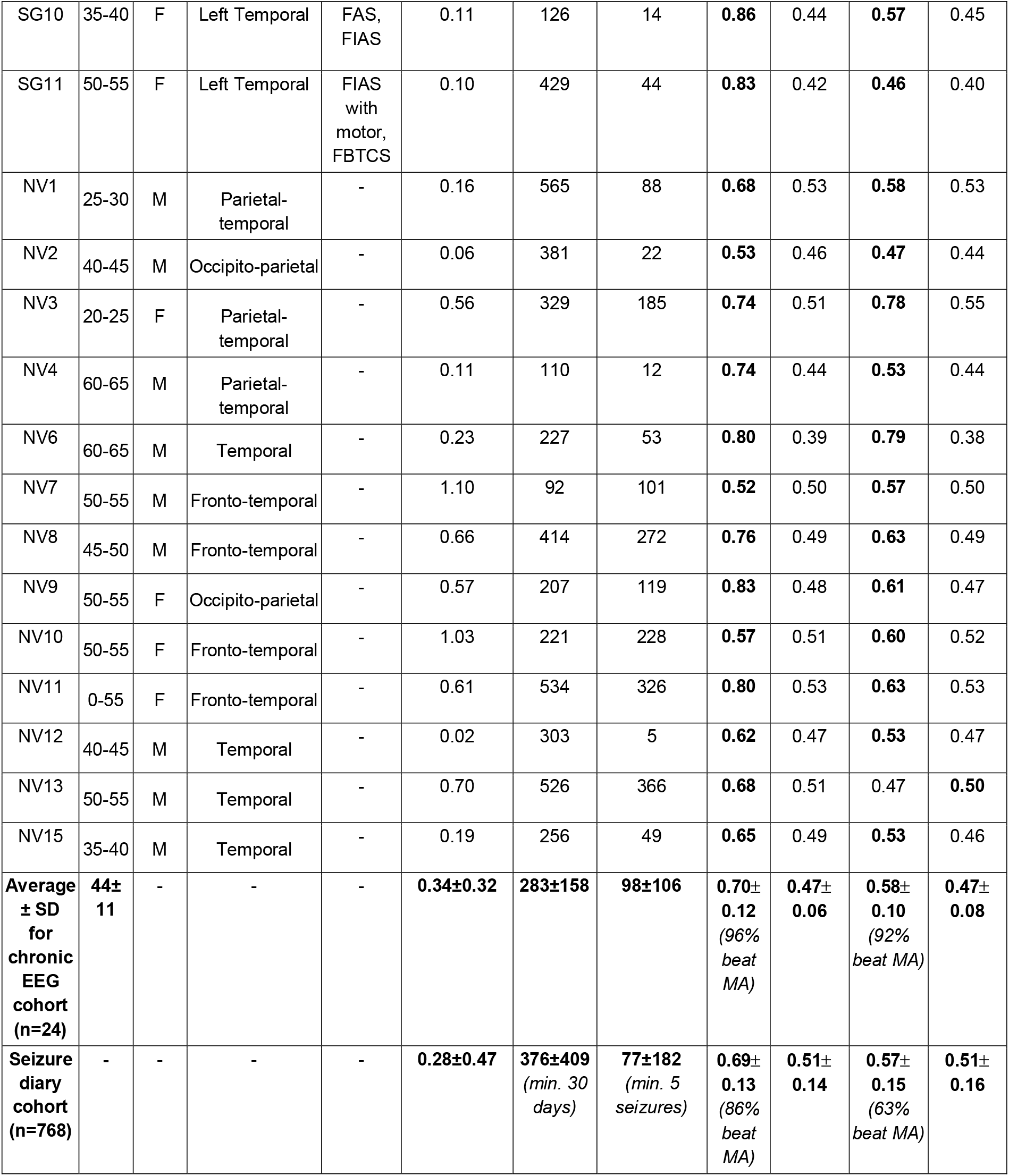
Subject demographic characteristics, clinical characteristics and pseudo-prospective forecasting results. SG = Seizure Gauge cohort, NV = NeuroVista cohort (participant IDs deidentified). FIAS = focal impaired awareness seizures, FAS = focal awareness seizures, TCS = tonic clonic seizures, FBTCS = focal to bilateral tonic clonic seizures Averages and standard deviations are reported for age, seizure frequency, and all pseudo-prospective results. Algorithms with superior performance (based on AUC) are bolded, per subject, at both hourly and daily timescales.

| Subject | Age | Sex | Localization | Seizure types | Seizure frequency during forecast period (per day) | Pseudo-prospective results |  |  |  |  |  |
| --- | --- | --- | --- | --- | --- | --- | --- | --- | --- | --- | --- |
|  |  |  |  |  |  | Forecast days | Forecasted seizures | Hourly cycles AUC | Hourly moving average AUC | Daily cycles AUC | Daily moving average AUC |
| SG1 | 40-45 | F | Bilateral Temporal | FIAS, FBTCS | 0.12 | 588 | 69 | **0.76** | 0.48 | **0.67** | 0.48 |
| SG2 | 35-40 | F | Left parietocentral and right frontocentral (L>R) | FIAS, FBTCS | 0.23 | 98 | 23 | **0.70** | 0.41 | **0.57** | 0.37 |
| SG3 | 50-55 | M | Independent Bitemporal | FAS, TCS | 0.59 | 269 | 159 | 0.53 | **0.63** | 0.49 | **0.67** |
| SG4 | 50-55 | M | Left Temporal | FBTCS | 0.08 | 226 | 19 | **0.90** | 0.40 | **0.48** | 0.41 |
| SG5 | 25-30 | M | Left Temporal | FIAS | 0.14 | 251 | 36 | **0.75** | 0.44 | **0.63** | 0.48 |
| SG6 | 40-45 | F | Bilateral Temporal | FIAS, FBTCS | 0.48 | 271 | 131 | **0.64** | 0.44 | **0.57** | 0.43 |
| SG7 | 40-45 | F | Left Frontocentral | FIAS | 0.15 | 78 | 12 | **0.70** | 0.44 | **0.78** | 0.49 |
| SG8 | 40-45 | M | Left Temporal | FIAS, FBTCS | 0.10 | 162 | 16 | **0.83** | 0.48 | **0.54** | 0.47 |
| SG9 | 20-25 | F | Left Temporal | FIAS, FBTCS | 0.04 | 121 | 5 | **0.49** | 0.30 | **0.37** | 0.27 |
| SG10 | 35-40 | F | Left Temporal | FAS, FIAS | 0.11 | 126 | 14 | **0.86** | 0.44 | **0.57** | 0.45 |
| SG11 | 50-55 | F | Left Temporal | FIAS with motor, FBTCs | 0.10 | 429 | 44 | **0.83** | 0.42 | **0.46** | 0.40 |
| NV1 | 25-30 | M | Parietal-temporal | - | 0.16 | 565 | 88 | **0.68** | 0.53 | **0.58** | 0.53 |
| NV2 | 40-45 | M | Occipito-parietal | - | 0.06 | 381 | 22 | **0.53** | 0.46 | **0.47** | 0.44 |
| NV3 | 20-25 | F | Parietal-temporal | - | 0.56 | 329 | 185 | **0.74** | 0.51 | **0.78** | 0.55 |
| NV4 | 60-65 | M | Parietal-temporal | - | 0.11 | 110 | 12 | **0.74** | 0.44 | **0.53** | 0.44 |
| NV6 | 60-65 | M | Temporal | - | 0.23 | 227 | 53 | **0.80** | 0.39 | **0.79** | 0.38 |
| NV7 | 50-55 | M | Fronto-temporal | - | 1.10 | 92 | 101 | **0.52** | 0.50 | **0.57** | 0.50 |
| NV8 | 45-50 | M | Fronto-temporal | - | 0.66 | 414 | 272 | **0.76** | 0.49 | **0.63** | 0.49 |
| NV9 | 50-55 | F | Occipito-parietal | - | 0.57 | 207 | 119 | **0.83** | 0.48 | **0.61** | 0.47 |
| NV10 | 50-55 | F | Fronto-temporal | - | 1.03 | 221 | 228 | **0.57** | 0.51 | **0.60** | 0.52 |
| NV11 | 0-55 | F | Fronto-temporal | - | 0.61 | 534 | 326 | **0.80** | 0.53 | **0.63** | 0.53 |
| NV12 | 40-45 | M | Temporal | - | 0.02 | 303 | 5 | **0.62** | 0.47 | **0.53** | 0.47 |
| NV13 | 50-55 | M | Temporal | - | 0.70 | 526 | 366 | **0.68** | 0.51 | 0.47 | **0.50** |
| NV15 | 35-40 | M | Temporal | - | 0.19 | 256 | 49 | **0.65** | 0.49 | **0.53** | 0.46 |
| Average ± SD for chronic EEG cohort (n=24) | 44±11 | - | - | - | **0.34±0.32** | **283±158** | **98±106** | **0.70±0.12**<br>(96% beat MA) | **0.47±0.06** | **0.58±0.10**<br>(92% beat MA) | **0.47±0.08** |
| Seizure diary cohort (n=768) | - | - | - | - | **0.28±0.47** | **376±409**<br>(min. 30 days) | **77±182**<br>(min. 5 seizures) | **0.69±0.13**<br>(86% beat MA) | **0.51±0.14** | **0.57±0.15**<br>(63% beat MA) | **0.51±0.16** |

The results shown in Table 1 show that event-based cyclical models tend to perform better compared to moving average when using EEG-confirmed seizures compared to self-reports. It is certainly true that any forecast derived from an inaccurate record of sself-reported events would not capture the underlying neurophysiological cycles and would perform poorly in a prospective or pseudoprospective setting. Given that half or more of epilepsy patients have been shown to keep inaccurate diaries, the low accuracy demonstrated by Chang et al and others (Elger & Hoppe, 2018; Goldenholz et al., 2024; Proix et al., 2021; Schulze-Bonhage et al., 2023; Wong et al., 2025) is not surprising.

However, this performance reflects a fundamental limitation of the data rather than any forecasting method. It is well established that self-reported seizure burden is not closely correlated with individuals’ electrographic seizure rates (Cook et al., 2013; Halliday et al., 2025; Wong et al., 2025). Nevertheless, for statistical reasons, a repeating pattern (i.e.a cycle) can be more reliably estimated from background noise; meaning that even unreliable self-reported seizures show similar periodic patterns as epileptic activity captured from EEG (Karoly et al., 2020; Leguia et al., 2021; Reynolds et al., 2025). This suggests that seizure cycles still provide a better *retrospective* biomarker of underlying changes in epileptic activity than self-reported seizure rates.

As more continuous data and reliable seizure detectors become available (either from wearable or implanted devices), the application of cycle-tracking becomes substantially more powerful, and it is unlikely that event-based moving average models will be able to match the performance of continuous cyclic forecasts. In any case, to suggest that a model is clinically useless because it can be computed ‘on the back of a napkin’ equates complexity with utility. We believe this is an error for medical applications, where ease of use, interpretability, and application should be prioritized. A forecasting method with many false alarms may cause alarm fatigue if sent to caregivers but may be helpful if integrated into a neuromodulation device. Further, most patients do not have an intuitive, everyday awareness of the current long-term trend in their seizure rates (the moving average model). Seizures are often rare, and humans are not naturally good at detecting historic health trends over months or years (Schmier & Halpern, 2004). Providing insight into past seizure patterns, whether a long-term trend or multiday cycle, is desirable to people with epilepsy (Grzeskowiak & Dumanis, 2021; Janse et al., 2019; Stirling et al., 2024; Goldenholz et al., 2024; Schmier & Halpern, 2004) and can drive improved quality of life (Stirling et al., 2024). It remains to be tested whether some people derive more benefit from a cycle vs a moving average model, and this likely depends on their individual statistical patterns and seizure frequency. Clearly, both models must be assessed, and the time has now arrived for these clinical trials.

## Data Availability

The data that support the findings of this study are available in Figshare (NV cohort seizure times) and Github (MSG cohort seizure times). Data from the seizure diary cohort can be provided upon reasonable request to the corresponding author. Code to compute the prospective moving average benchmark is available from Github at https://github.com/RiPLresearch/forecast-benchmarks/tree/main and code to compute prospective seizure cycles can be shared upon reasonable request to the corresponding author.

https://www.epilepsyecosystem.org/my-seizure-gauge-1

https://doi.org/10.26188/5b6a999fa2316

